# Investigating the use and effectiveness of lifestyle medicine approaches to tackle erectile dysfunction: a cross-sectional eSurvey based study

**DOI:** 10.1101/2022.05.30.22275800

**Authors:** Austen El-Osta, Gabriele Kerr, Aos Alaa, Marie Line El Asmar, Manisha Karki, Iman Webber, Eva Riboli Sasco, Giordano Blume, Wolf-D. Beecken, David Mummery

## Abstract

**Background:** Erectile dysfunction (ED) is the most common sexual dysfunction in men. Some types of ED are amenable to treatment using lifestyle medicine approaches with or without pharmacotherapy.

**Aim:** Investigate the use and perceived effectiveness of lifestyle medicine approaches to tackle ED.

**Methods:** A cross-sectional online survey of 1177 community dwelling adults explored the prevalence and methods used to tackle ED in the community setting. We examined differences between participants with and without ED. Variables associated with ED in univariable analyses were included in a multivariable logistic regression to identify variables independently associated with the condition.

**Outcomes:** Self-reported measure; perceived effectiveness of lifestyle medicine interventions to tackle ED

**Results:** Most respondents (76.5%) had experienced ED, and this was associated with having a long-term condition, taking anti-hypertensive medication, hypercholesterolaemia and obesity. Medication was the most common management strategy overall (65.9%), followed by stress management (43.5%) and weight loss (40.4%). Over half (53.9%) did not use any lifestyle modification strategies to tackle ED. Only 7.0% of ED sufferers received a mental health assessment and 29.2% received other tests (e.g., blood test, medical imaging) by GPs. Cardiovascular training was identified as the best rated strategy by its users (37.8%). Supplements (35.1%) and weight training/physical activity (32.6%) were also positively rated.

**Clinical implications:** Structured education to general practitioners and community dwelling adults about the impact of lifestyle behaviour modification and how this could influence the appearance or trajectory of ED could help improve personal choice when tackling ED.

**Strengths and Limitations:** To our knowledge, this is the first study to collect eSurvey responses from community dwelling adults to gauge their reliance and perceived effectiveness of lifestyle medicine approaches to tackle ED. The principal limitation was the lack of follow-up, and not recording other information including lifestyle factors such as nutrition, smoking, and the use of alcohol and recreational drugs, which may have enabled a fuller exploration of the factors that could influence the primary outcome measures examined.

**Conclusion:** Despite the high prevalence of ED, there is not enough awareness in the community setting about effective and low-cost lifestyle medicine strategies, including cardiovascular training and the use of supplements and weight training, to help tackle this common condition

## Introduction

Erectile dysfunction (ED) refers to the inability to achieve or maintain a rigid penile erection (1) and is the most common sexual dysfunction in men, followed by decreased libido and ejaculatory dysfunction, which can co-exist simultaneously (2). There are no uniform criteria that define how consistent the problem needs to be to qualify for a diagnosis of ED, but usually indicated when the dysfunction lasts six months or longer (3, 4).

ED has a high reported prevalence in the general population and affects up to a third of men worldwide and an estimated one in five (4.3 million) men in the UK (5-14). These figures likely underestimate the actual prevalence of ED due to reporting bias, cultural factors, physicians not inquiring about the patients’ sexual health, and the stigma associated with the dysfunction (15). Figures show a discrepancy, with prevalence higher in the United States and south-eastern Asian countries than in Europe and South America, likely due to cultural and socioeconomic factors, but genetic factors may also play a role (7, 15-17). By 2025, the prevalence is estimated to reach approximately 322 million men worldwide (14). Individuals with chronic systemic disorders such as hypertension, ischemic heart disease, and diabetes mellitus are more likely to develop ED, and its occurrence rises steeply in men aged 40 years or over (18).

ED can have multiple underlying factors including; cardiovascular (diabetes mellitus, hyperlipidemia, hypertension, metabolic syndrome and obesity), neurologic or psychological factors (stress, anxiety, depression, body image, self-esteem, performance anxiety, and relationship issues), hormonal (testosterone deficiency, or thyroid disorders), traumatic (genitourinary trauma, spinal cord injuries), and medication and substance use including antidepressants, antihypertensives, antipsychotics, opioids, tobacco, and recreational drugs (19-27). ED often precedes the onset of cardiovascular disease (CVD) and may be considered an indicator or an early marker from a clinical perspective (28). While the condition is more common in the older population, younger men aged 18 to 25 years can also be affected, usually with a psychogenic rather than organic underlying cause (29).

Conventional medical therapy includes the use of phosphodiesterase-5 inhibitors (PDE5I) as first-line treatments for ED (19). Other treatments include penile self-injections with vasoactive drugs and intraurethral suppositories (30, 31), and when other therapeutic treatment options fail to yield results, surgically implanted penile prostheses may be considered (19). Alongside medical treatment, lifestyle and behaviour modifications such as weight management, uptake of physical activity, smoking and alcohol cessation, improving blood glucose, hyperlipidaemia and blood pressure control are usually recommended as initial interventions by guidelines (32).

Lifestyle changes can significantly improve sexual function in some individuals experiencing ED (33-39). Observational studies highlight a link between smoking and alcohol consumption (40), and high-fat, high sugar diets (41, 42) and ED, whereas conversely exercising ≥18 metabolic equivalent hours/week was associated with improved sexual function (43). Another study showed that men who underwent gastric bypass surgery, which usually results in significant weight loss, had improved testosterone levels and erectile function (44). These data suggest that self-care using behaviour and lifestyle modification to control these modifiable factors may prevent, delay the appearance of ED, or alter the trajectory of or enhance the regression of symptoms and manifestations of ED (45). In this regard, modifiable risk factors including tobacco and alcohol use, weight management and physical activity are a key area of focus when managing ED (36, 37, 46-49).

There were previous attempts to investigate opinions and attitudes regarding ED, and its effect on the quality of life and on masculinity (50-53) but public perception of the value of lifestyle modifications in managing ED remain largely understudied. Considering that pharmacotherapy is currently the most common treatment modality for ED, which is rising in prevalence, and that lifestyle modification is associated with significant improvement of ED, understanding the update and perceived benefits of lifestyle interventions in managing ED is essential. The aim of our study was to investigate the use and perceived effectiveness of using of lifestyle medicine approaches to tackle erectile dysfunction.

## Methods

### Study design and setting

A community-based cross-sectional study was conducted using an electronic survey administered on Qualtrics and distributed through social media outlets.

### Data collection

Data from 1177 survey respondents was collected using a short (<10 min) anonymised electronic survey using convenient sampling. The survey link was made available on the Imperial College London Qualtrics platform for 17 weeks between Nov 21 and Feb 22.

Participants were also able to participate through community group distribution lists, social media advertisements including on Reddit, Twitter, and other outlets. The eSurvey was accessible by anyone with a link via a personal computer or smartphone (open survey). Information about the study, including the Participant Information Sheet (PIS), was provided in the introductory section of the survey. The PIS detailed the study’s objectives, protecting participants’ data, their right to withdraw at any stage, information on where, when, and for how long data were retained, the study’s investigator, and the survey’s length. There were no monetary incentives offered to participants for completing this voluntary survey, but they were informed that their participation could help advance our knowledge about how lifestyle modification may be used as a method to tackle ED and were offered the possibility to access research findings. Survey results were stored on Imperial College London’s secure database, which could only be accessed by the research team.

The Qualtrics survey (Version XM) contained 30 questions all on one page and automatically captured responses. Qualtrics’s websites have first party cookies and allow third parties to place cookies on devices. The survey’s technical functionality and usability was developed and piloted on six researchers before online dissemination. The survey was published in 10 languages (English, Arabic, German, Polish, Punjabi, Somali, Urdu, Persian, French & Spanish), involved 27 questions of tick or Likert-scale responses which could be answered using a personal computer or smartphone. The survey included conditional questions that would appear depending on which options they selected. Questions were designed to gauge the knowledge, attitudes, and perceptions of respondents regarding ED interventions and to better understand the extent that lifestyle medicine approaches were being used to tackle ED in the community setting. Study participants included individuals 18 years or over but excluded those with no access to the internet, or who did not provide consent. Participants could review their answers before submitting them. No IP addresses were collected; therefore, the team could not identify any cases of duplicate entries. Survey responses were only excluded if most of the survey was incomplete.

Initially, the survey sought to understand respondents’ past experiences with erections, including whether such difficulty lasted for 3-6 months, their confidence in getting and maintaining an erection over the past 3-6 months, and how vital having a firm erection is to their quality of life and wellbeing. Respondents who reported difficulty with erections were asked how they resolved their issue, if they sought professional help, what advice they received (whether through medication, lifestyle changes, or other means), and whether it helped them improve the quality of erections. Additionally, the survey asked respondents what factors they think cause ED, what approaches can alleviate ED, and what measures (including lifestyle changes) they have previously tried to improve the quality of their erection. Respondents were then asked if they would consider using lifestyle medicine as a standalone therapy or as an adjunct to other treatments to improve erection quality. The survey ended with routine lifestyle, health and demographic questions including gender, age and ethnicity. Consent for participation was taken, and collected data were anonymized. The complete survey is accessible using this link: https://imperial.eu.qualtrics.com/jfe/form/SV_b715ofTQigWYqPk. The quality of the survey was assessed using the Checklist for Reporting Results of Internet E-Surveys (CHERRIES).

### Data analysis

Participants who stated they had issues in getting an erection over an extended period (3-6 months) were considered to have ED. Participant characteristics were summarized for respondents with and without ED using descriptive statistics using frequencies and percentages for categorical data and median with interquartile range (IQR) for quantitative data. Pearson’s χ^2^ test and Mood’s median test were used to examine differences between participants with and without ED. An α level of significance of 0.05 was used throughout. Variables associated with ED in univariable analyses were included in a multivariable logistic regression to identify variables independently associated with ED. Odds ratios (OR) and 95% confidence intervals (CI) were calculated. Analyses were conducted in R version 4.1.2.

### Ethics approval

The study received a favourable opinion from Imperial College Research Ethics Committee (21IC7318). Participants consented to take part in the survey.

### Patient and Public Involvement

No patient was involved.

## Results

The electronic survey received 1177 total responses. Only 490 participants (41.6%) remained after excluding incomplete surveys.

The sociodemographic and health characteristics of the survey respondents by ED status are shown in **table 1**. The study population was 98% male, 91.4% of white ethnicity, and had a median age of 63 (IQR 53 – 70) years, with a range of 18 to 85 years. Most (76.5%) participants had experienced issues with getting an erection over an extended period and were therefore classed as having ED. Almost half (45.1%) of respondents were taking either an antidepressant, anti-hypertensive, or anti-androgenic medication. Most (70.0%) were in a relationship and 72.2% were sexually active, 90.6% were not regular smokers, 42.2% reported being overweight, 60.4% described their diet as very or generally healthy, and 79.4% exercised at least once a week; **table 1**. One third (33.2%) of participants reported having a mental health condition and 12.4% stated they had a disability. Of the 43.3% who reported having a long-term condition, most had high blood pressure, with 20.4% of participants overall stating they had the condition.

**Table 1:** Characteristics of survey respondents by ED status (N=490)

|  | No ED (n = 115)<br>N (%) | ED (n = 375)<br>N (%) | p-value* |
| --- | --- | --- | --- |
| **Gender** |  |  | 0.21 |
| Female | 2 (1.7) | 1 (0.3) |  |
| Male | 112 (97.4) | 368 (98.1) |  |
| Other | 1 (0.9) | 2 (0.5) |  |
| **Age** |  |  | <0.001 |
| Median (IQR) | 56 (38.0 – 65.0) | 64 (57.0 – 70.0) |  |
| **Relationship Status** |  |  | 0.33 |
| Divorced | 6 (5.2) | 24 (6.4) |  |
| Married | 61 (53.1) | 215 (57.3) |  |
| Single | 21 (18.3) | 72 (19.2) |  |
| In a domestic relationship | 22 (19.1) | 45 (12.0) |  |
| Widowed | 2 (1.7) | 14 (3.7) |  |
| Other | 3 (2.6) | 5 (1.3) |  |
| **Ethnicity** |  |  | 0.31 |
| Asian/Asian British | 6 (5.2) | 7 (1.9) |  |
| British Black/ African/ Caribbean | 0 (0.0) | 4 (1.1) |  |
| Mixed/Multiple ethnic groups | 2 (1.7) | 12 (3.2) |  |
| White | 105 (91.3) | 343 (91.7) |  |
| White & Black Caribbean | 0 (0.0) | 1 (0.3) |  |
| Other | 2 (1.7) | 8 (2.1) |  |
| **Sexually active** |  |  | <0.001 |
| No | 16 (13.9) | 120 (32.0) |  |
| Yes | 99 (86.1) | 255 (68.0) |  |
| **Regular Smoker** |  |  | 0.44 |
| No | 106 (93.0) | 338 (90.6) |  |
| Yes | 8 (7.0) | 35 (9.4) |  |
| **Regular Smoker** |  |  | 0.24 |
| Never | 12 (10.4) | 24 (6.4) |  |
| Less than once a week | 12 (10.4) | 53 (14.1) |  |
| Once a week | 12 (10.4) | 56 (14.9) |  |
| 2-3 times a week | 50 (43.5) | 137 (36.5) |  |
| 4 or more times a week | 29 (25.2) | 105 (28.0) |  |
| **Overweight** |  |  | 0.04 |
| No | 76 (66.1) | 206 (55.2) |  |
| Yes | 39 (33.9) | 168 (45.0) |  |
| **Diet** |  |  | 0.25 |
| Very healthy | 7 (6.1) | 37 (9.9) |  |
| Generally healthy | 64 (55.7) | 188 (50.1) |  |
| Average | 38 (33.0) | 130 (34.7) |  |
| Generally unhealthy | 5 (4.4) | 20 (5.3) |  |
| Very unhealthy | 1 (0.9) | 0 (0.0) |  |
| **Disability** |  |  | 0.16 |
| No | 104 (90.4) | 303 (81.2) |  |
| Yes | 10 (8.7) | 51 (13.7) |  |
| **Mental health condition** |  |  | 0.46 |
| No | 79 (68.7) | 248 (66.5) |  |
| Yes - Depression | 12 (10.4) | 63 (16.9) |  |
| Yes - anxiety | 14 (12.2) | 42 (11.3) |  |
| Other condition | 3 (2.6) | 8 (2.1) |  |
| **Long-term condition** |  |  | 0.001 |
| No | 77 (67.0) | 184 (49.3) |  |
| Yes | 35 (30.4) | 177 (47.5) |  |
| **Specific long-term condition** |  |  |  |
| High blood pressure | 14 (12.2) | 86 (22.9) | 0.35 |
| Cardiovascular disease | 3 (2.6) | 36 (9.6) | 0.10 |
| Diabetes mellitus | 7 (6.1) | 45 (12.0) | 0.50 |
| High blood cholesterol | 4 (3.5) | 51 (13.6) | 0.03 |
| Other | 20 (17.4) | 66 (17.6) | 0.03 |
| **Taking anti- depressant, hypertensive, or androgenic medication** |  |  | <0.001 |
| No | 80 (69.6) | 173 (46.1) |  |
| Yes | 35 (30.4) | 202 (53.9) |  |
| **Specific Medication** |  |  |  |
| Antidepressant | 13 (11.3) | 65 (17.3) | 0.16 |
| Anti-hypertensive | 23 (20.0) | 148 (39.5) | <0.001 |
| Anti-androgenic | 3 (2.6) | 23 (6.1) | 0.22 |

\* Comparison of participants with and without ED. | Data shown as n (%) for categorical data and median (IQR) for continuous data. Percentages may not add up to 100% as numbers for “unknown” categories are not shown here. Statistical tests are $\chi^2$ test for categorical variables and Mood’s median test for continuous variables. Abbreviations: ED – erectile dysfunction; IQR – interquartile range; n – number.

In univariable analyses, there was no evidence for differences in smoking or exercise habits, diet, relationship status, ethnicity, or likelihood of having mental health conditions between respondents who have or have not had ED. Compared to participants without ED, those who experienced ED in the last 3-6 months more often reported having a long-term condition, taking anti-hypertensive medication, having high were typically and less frequently reported being sexually active. The following variables were therefore included in the multivariable logistic regression model: age, taking antihypertensive medication, being overweight, having a long-term condition, having high blood cholesterol, and being sexually active. In the multivariable regression analysis, only two variables were independently associated with ED status and were retained in the final model: age (OR: 1.04, 95% CI; 1.021 - 1.05, p < 0.001) and sexual activity (OR: 0.38, 95% CI; 0.21 - 0.66, p = 0.001).

### Prevalence of ED

Most participants (83.7%) reported ever having had an experience where they could not get an erection, with 76.5% stating they had experienced issues with getting an erection over an extended period. When asked about their confidence to get or to maintain an erection over the last 3-6 months, 48.6% and 53.5% of participants responded ‘low’ or ‘very low’ in that same order. Respondents with ED more often reported having lower confidence in both regards compared to participants without ED (**table 2**; p<0.001).

**Table 2:**
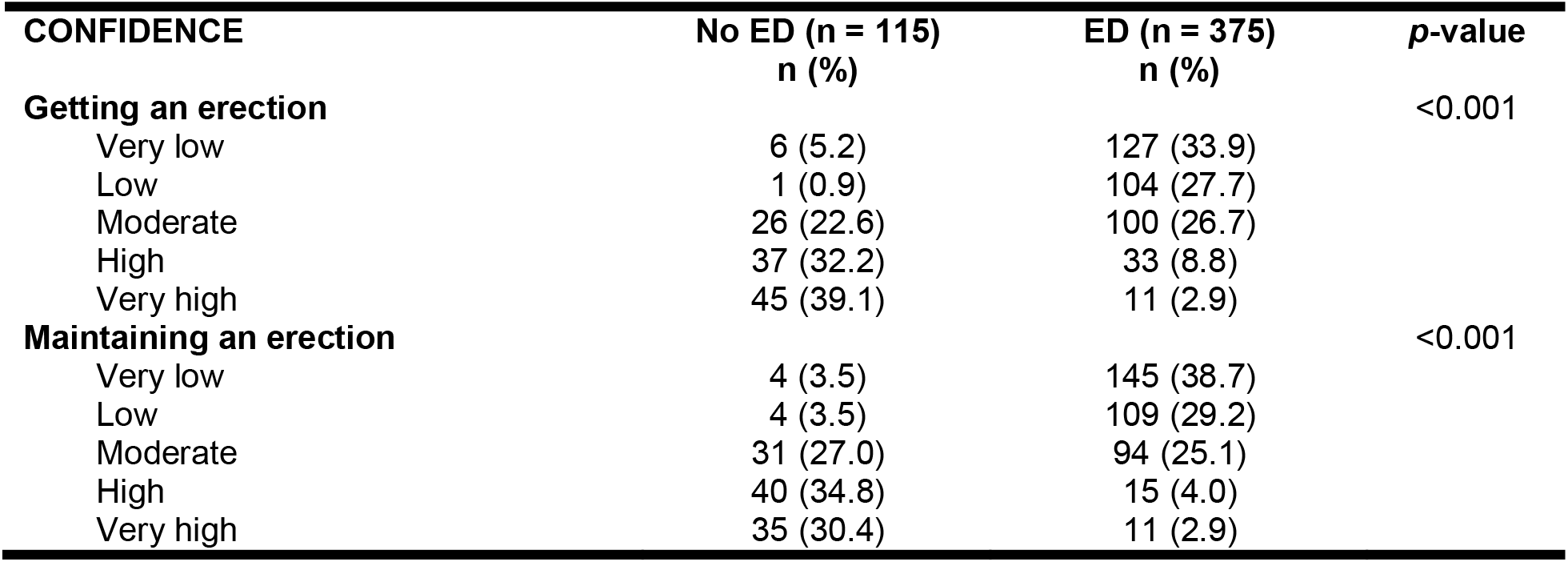
Participant confidence in their ability to get or maintain an erection by ED status.

| CONFIDENCE | No ED (n = 115)<br>n (%) | ED (n = 375)<br>n (%) | p-value |
| --- | --- | --- | --- |
| **Getting an erection** |  |  | <0.001 |
| Very low | 6 (5.2) | 127 (33.9) |  |
| Low | 1 (0.9) | 104 (27.7) |  |
| Moderate | 26 (22.6) | 100 (26.7) |  |
| High | 37 (32.2) | 33 (8.8) |  |
| Very high | 45 (39.1) | 11 (2.9) |  |
| **Maintaining an erection** |  |  | <0.001 |
| Very low | 4 (3.5) | 145 (38.7) |  |
| Low | 4 (3.5) | 109 (29.2) |  |
| Moderate | 31 (27.0) | 94 (25.1) |  |
| High | 40 (34.8) | 15 (4.0) |  |
| Very high | 35 (30.4) | 11 (2.9) |  |

Overall, 81.8% of participants stated that a firm and lasting erection was “important” or “very important” for their mental and physical wellbeing and quality of life. Nearly half (46.4%) of participants with ED stated that a firm and lasting erection was “very important” to their health and wellbeing compared to only 26.9% of participants without ED.

### Discussing ED

Most participants (75.1%) stated they would feel comfortable discussing ED with a GP or doctor, sexual health clinic staff (63.5%), or partner/spouse (58.8%). Fewer participants were comfortable discussing the topic with pharmacists (30.0%), online chatbots (23.5%), friends (22.9%) or family (19.4%). A minority (8.0%) did not feel comfortable discussing ED with any of the suggested options. Responses between the two groups were very similar, except that those with ED more often stated they would be comfortable discussing ED with a pharmacist (p<0.001) but less frequently stated they would be comfortable discussing ED with a partner or spouse (p=0.02).

### How and where people receive information about ED

Nearly half of participants with ED (45.6%) sought information about tackling the dysfunction from a GP/health care professional (HCP); 28.0% had sought information only from a HCP and no other source. Of the 171 participants that sought information about resolving ED from HCP, 73.7% were offered advice in the form of medication, whereas only 12.3% received advice about lifestyle changes. The majority (63.7%) of these participants reported that the HCP did not run any tests to identify the cause of ED. Only 7.0% received a mental health assessment, and 29.2% received other tests (e.g., blood tests, medical imaging). Many respondents used online websites (39.2%) or online forums (19.7%), with 29.9% stating they sought information from online websites or forums and no other sources. Very few (3.2%) participants with ED sought information from friends or family.

### Causes of ED

More than half (61.0%) of respondents felt that ‘stress or poor mental health’ was the most frequent cause of ED, although this option was less frequently selected by respondents with ED than by those without the condition **(table 3; figure 1)**. Other factors, including smoking, unhealthy diet, obesity, certain medications, alcohol or other substance abuse, and previous pelvic surgeries or spinal injuries, were more often identified as causes of ED by respondents without ED than by those with ED.

**Table 3:** Participant identified causes of ED.

| Perceived cause of ED | No ED (n = 115)<br>n (%) | ED (n = 375)<br>n (%) | P value |
| --- | --- | --- | --- |
| Stress/other mental health conditions | 101 (87.8) | 198 (52.8) | <0.001 |
| Unhealthy diet | 57 (49.6) | 116 (30.9) | <0.001 |
| Smoking | 54 (46.9) | 108 (28.8) | <0.001 |
| Obesity | 61 (53.0) | 144 (38.4) | 0.005 |
| Certain medications | 74 (64.4) | 192 (51.2) | 0.01 |
| Health conditions | 70 (60.9) | 208 (55.5) | 0.31 |
| Alcohol or other substance abuse | 85 (73.9) | 139 (37.1) | <0.001 |
| Previous pelvic surgeries or spinal injuries | 57 (49.6) | 106 (28.3) | <0.001 |
| Other | 15 (13.0) | 73 (19.5) | 0.12 |

**Figure 1:**
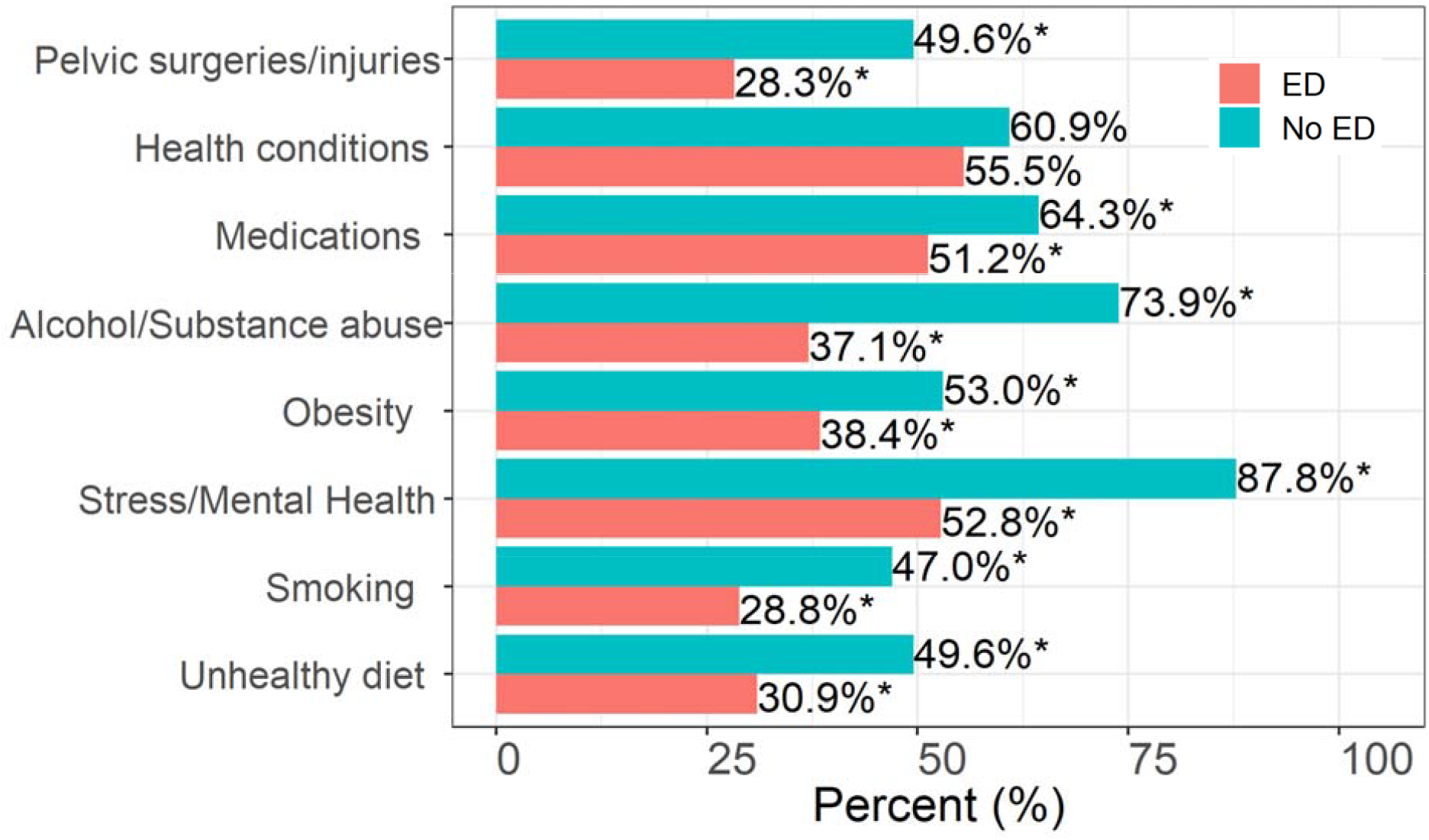
Participant perceived causes of ED, by ED status. An asterisk indicates significance at *p*<0.05.

### Management of ED

Respondents were asked to select which options from a given list were potential management strategies for ED (**table 4; figure 2)**. Medication was the most frequently identified management strategy overall (65.9%), followed by stress management (43.5%) and weight loss (40.4%). Supplements (14.5%), vacuum pump training (19.2%) and pelvic floor training (22.9%) were the least frequently selected. One third (34.7%) of participants did not identify any lifestyle modification approaches as potentially beneficial for the management of ED. There were differences in the selection of potential management strategies between participants with and without ED. The former identified fewer approaches than participants without ED (*p*<0.001), whereas participants with ED identified a median of 3 (IQR 1 – 5) approaches, whereas participants without ED selected a median of five approaches (IQR 2 – 8). Only vacuum pump training and supplements were more commonly selected by participants with ED than by those without **(table 4)**. There were no differences in selection frequencies between the two groups for testosterone replacement therapy and medication. All other options were more commonly chosen by participants without ED than by those with ED.

**Table 4:**
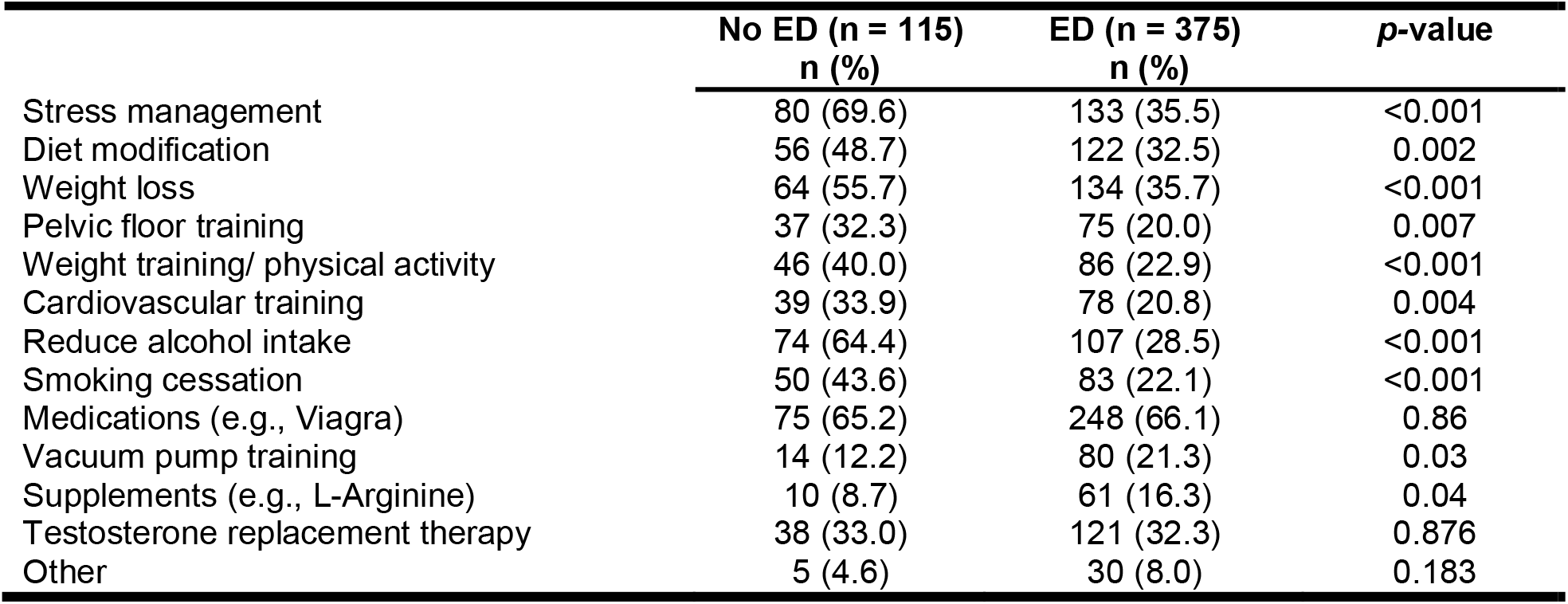
Participant identified approaches for management of ED, by ED status.

**Figure 2:**
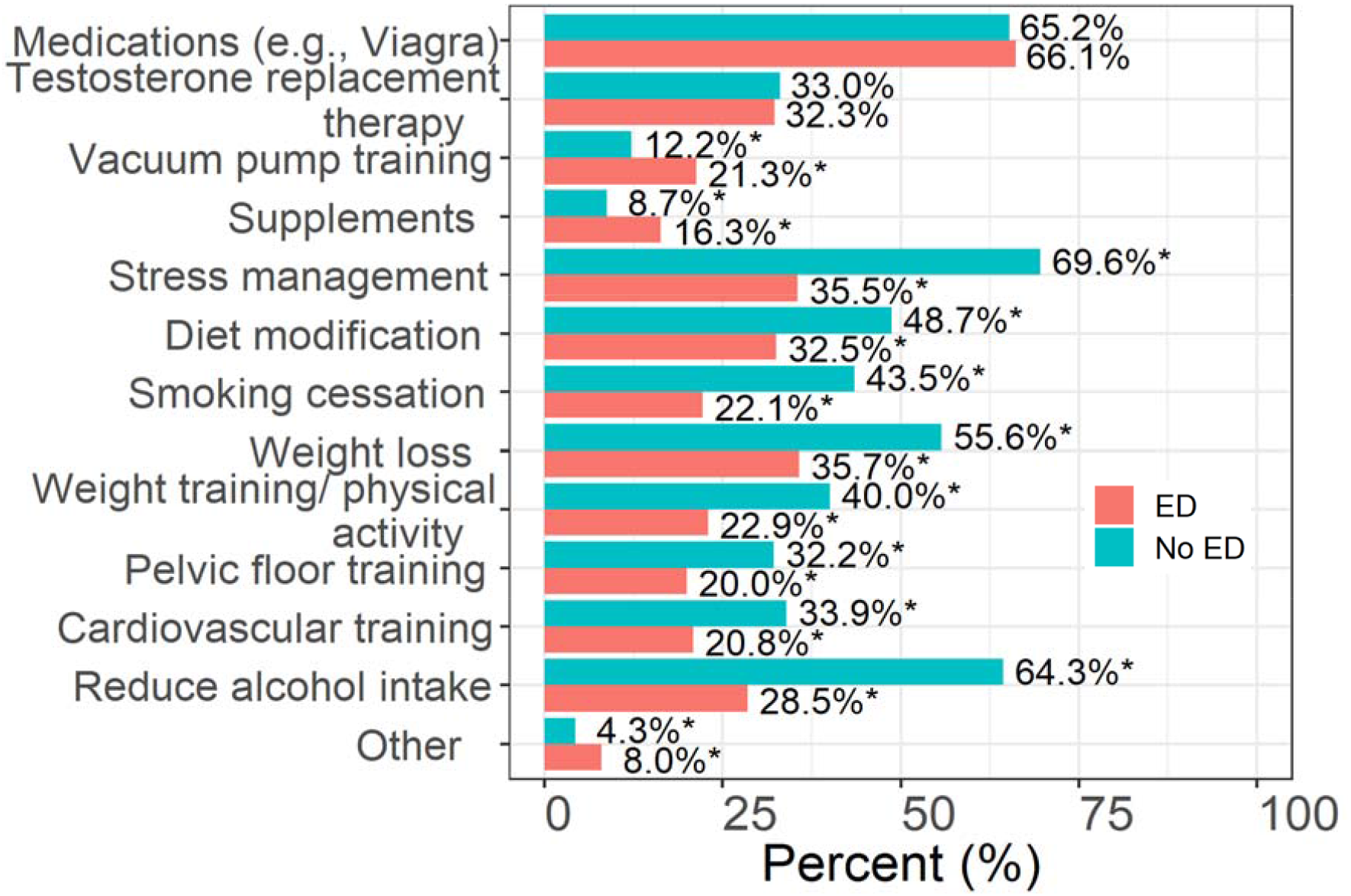
Participant reported effective approaches for the management of ED, by ED status. An asterisk indicates significance at *p*<0.05.

### Strategies to tackle ED

Strategies personally employed by participants with ED to alleviate the condition are described in **table 5**. Nearly one third (32.3%) of participants selected only one strategy. Over half (54.9%) used medications to alleviate ED, with 20.8% having only used medications and no other strategy. Over half (53.9%) had not used any lifestyle modification strategies.

**Table 5:** Management strategies employed by respondents with ED and their effect on erection quality as rated by users (n= 375)

| Management strategy | Total<br>N (% of 375) | Rating, n (% of total) |  |  |
| --- | --- | --- | --- | --- |
|  |  | No effect | Moderate effect | Significant effect |
| Stress management | 74 (19.7) | 23 (31.1) | 32 (43.2) | 19 (25.7) |
| Diet modification | 68 (18.1) | 23 (33.8) | 33 (48.5) | 12 (17.7) |
| Weight loss | 62 (16.5) | 22 (35.5) | 28 (45.2) | 12 (19.4) |
| Pelvic floor training | 43 (11.5) | 13 (30.2) | 19 (44.2) | 11 (25.6) |
| Weight training/ physical activity | 46 (9.9) | 9 (19.6) | 22 (47.8) | 15 (32.6) |
| Cardiovascular training | 37 (9.9) | 10 (27.0) | 13 (35.1) | 14 (37.8) |
| Reduce alcohol intake | 61 (16.3) | 19 (31.2) | 32 (52.5) | 9 (14.8) |
| Smoking cessation | 25 (6.7) | 8 (32.0) | 14 (56.0) | 3 (12.0) |
| Medications (e.g., Viagra) | 206 (54.9) | 42 (20.4) | 102 (49.5) | 61 (29.6) |
| Vacuum pump training | 36 (9.6) | 14 (38.9) | 16 (44.4) | 6 (16.7) |
| Supplements (e.g., L-Arginine) | 37 (9.9) | 10 (27.0) | 14 (37.8) | 13 (35.1) |
| Testosterone replacement therapy | 21 (5.6) | 7 (33.3) | 7 (33.3) | 7 (33.3) |

Although only 9.9% of participants had employed cardiovascular training, it was the best rated strategy by its users overall, with 37.8% describing it as having a significant effect on the quality of their erection. Supplements and weight training/physical activity were also positively rated, with 35.1% and 32.6% of their respective users describing them as having a significant effect. The worst rated strategy by users was vacuum pump training, with 38.9% of users reporting it had no effect on erection quality.

Overall, most respondents with ED stated they would consider using a lifestyle medicine app as an adjunct to other treatments (42.1%) or standalone (20.8%), to prevent, manage, or help improve the quality of their erections.

## Discussion

To our knowledge, this was the first study that sought to investigate public knowledge, attitudes, and perceptions about using lifestyle medicine approaches to tackle erectile dysfunction. Our sample largely consisted of respondents who have or are currently still experiencing ED (76%) but this was expected given the high prevalence of this condition with increasing age, and the special interest that individuals suffering from ED would have in a research study about this personal issue which directly impacts the quality of their daily life.

We found significant differences in the knowledge & perceptions of individuals affected by ED when compared to other members of the community who did not experience the condition. A key finding was that whereas nearly half of those affected by ED sought help from their GP, only a third of patients received further investigations to understand the root cause of the dysfunction. This was in spite current best practice guidelines and the availability of routine investigations in primary care (**table 6**). Another key finding was that GPs far more often recommend medication than lifestyle advice to patients seeking support, regardless of whether further investigations were made, such that only 12% were recommended a change in lifestyle. This highlights a problem of communication between the physician and the patients when addressing this common condition.

**Table 6:**
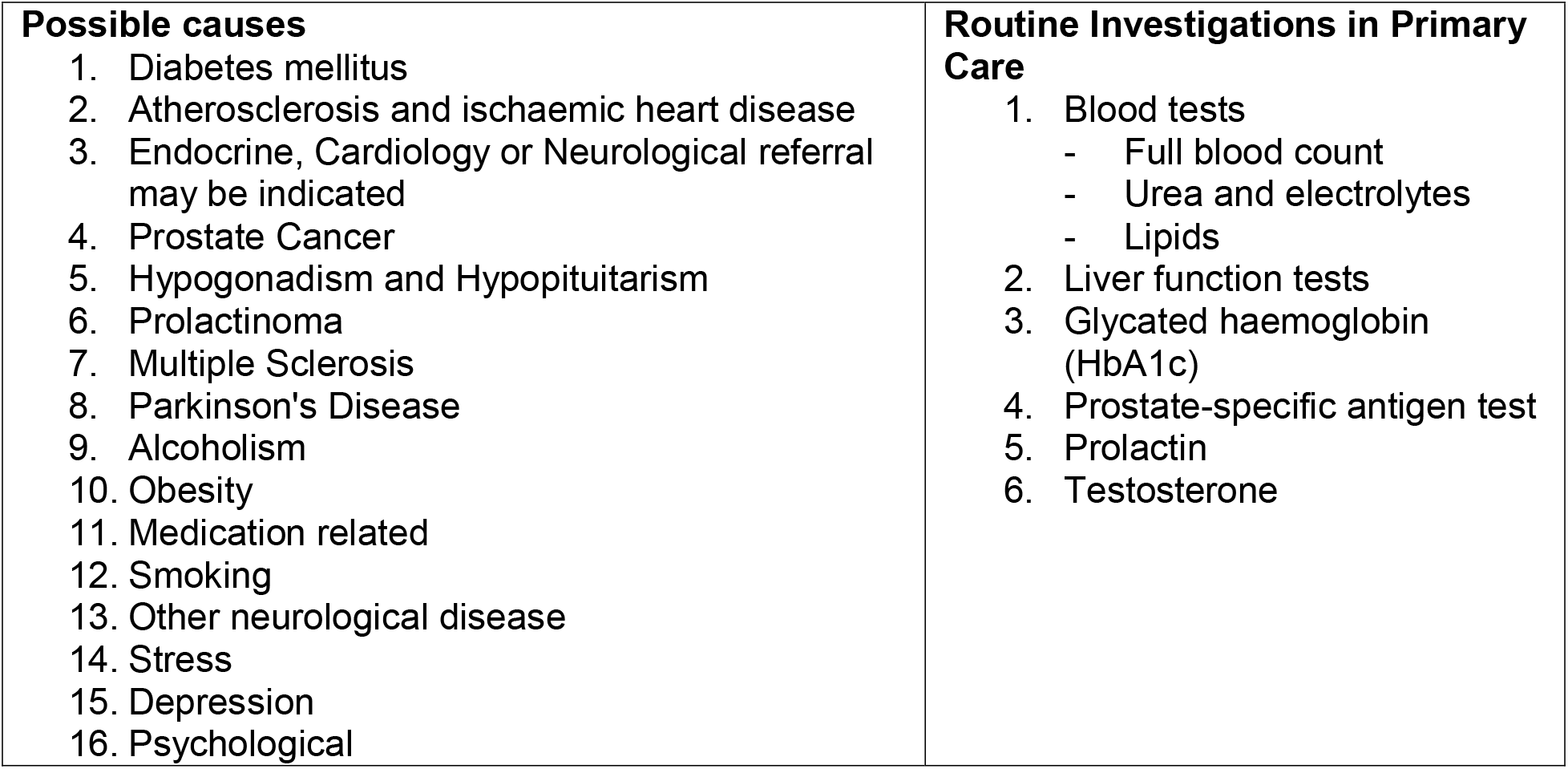
Possible causes of ED and Routine Investigations in Primary Care.

Another key finding was the reported effectiveness of cardiovascular training. Despite the small and cross-sectional nature of the sample, this lifestyle intervention was ranked the highest, even above the use of conventional pharmacotherapy (**table 5**). In this context, the reported value of other behaviours such as weight training and physical activity can also be considered as additional support for the routine adoption of lifestyle medicine approaches to tackle ED.

Our community survey showed that 72.9% of respondents with ED said that cardiovascular training had a moderate or significant effect on erection quality, whereas 80.4% of respondents reported weight training/ physical activity had the same effect. In comparison, 79.1% reported a moderate or significant effect on erection quality with the aid of medication. This shows that a lifestyle medicine approach can be as effective, if not more so, than medications such as sildenafil or tadalafil. These findings highlight a major problem in the care of patients with erectile dysfunction in that only half of patients have ever tried any lifestyle change, although the majority (75%) of patients who used lifestyle modification reported a moderate or significant effect of physical activity (e.g., cardiovascular training, weight training)

### Study Implications

GPs are generally regarded as the most common trusted source of information and help, but the received treatment for ED in the primary care setting is in most cases not fulfilling and lacks medical checks as recommended by current best practice guidelines. In view of this, the increased interest in lifestyle medicine apps solutions that promote adherence to key behaviour lifestyle modifications highlights the eagerness of ED sufferers to feel empowered and self-care for their condition. The ability of an individual to self-care is dependent on many factors (54) but can be supported by the rational use of ‘digital pill’ solutions with added functionality to educate patients about the root cause of their condition whilst providing evidence-based recommendations for positive behaviour change and lifestyle modification.

The very short (8 min) average consultation time with a GP does not allow for a lengthy communication about the importance of lifestyle changes and the sustained adoption of health-seeking self-care behaviours, especially for conditions like ED where pharmacotherapy is usually a convenient option. However, since medication alone is not described as more effective by affected patients and the sustained on-demand use of PDE5I does not improve or resolve the root cause of the dysfunction, patients should be more routinely given access to information and guidance for lifestyle medicine approaches to tackle ED. This approach is recommended as more than 60% of ED sufferers surveyed in this study reported they would consider using a lifestyle medicine app to tackle ED with or without pharmacotherapy and/or to improve the quality of their erection.

Given the pervasive use of smartphones, well-designed health apps that offer a digital pill solution to common conditions including ED could help with personal empowerment, improved health literacy and the sustained adherence to health-seeking self-care behaviours to prevent, delay the appearance of or improve the trajectory of ED. Further, since ED is a known early warning signal for cardiovascular events, greater awareness about the causes of ED coupled to timely lifestyle adjustment in the community setting could result to the earlier identification of at-risk patient groups.

### Strengths and limitations

The principal limitation of our study was the lack of follow-up, and not recording information about household income and other demographic, digital literacy, and lifestyle factors such as nutrition, smoking, and the use of alcohol and recreational drugs, which may have enabled a fuller exploration of the factors that could influence the primary outcome measures examined. Further, the demographic profile of study participants largely consisted of white men in full or part-time employment, implying that this cross-sectional study may not be representative of the wider UK population. We also acknowledge that since this was an online survey, we may have excluded individuals with limited digital access, and this restricts the generalisability of our findings to the wider population. Despite these limitations, our study highlights key findings that could be explored further using a personal interview component with men who are experiencing ED, healthy volunteers who did not suffer from this condition and healthcare professionals, including GPs to further explore barriers and drivers for the routine adoption of lifestyle medicine approaches to tackle ED.

### Conclusion

Despite the high prevalence of ED, there is not enough awareness in the community setting about effective and low-cost lifestyle medicine strategies, including cardiovascular training and the use of supplements and weight training, to help tackle this common condition. Structured education about the impact of behaviour and lifestyle modification and how this could influence the appearance or trajectory of ED could help improve personal choice and empowerment. A larger prospective cohort study or a randomised controlled trial is indicated to shed light on the effectiveness of lifestyle medicine approaches to tackle ED in the community setting with or without pharmacotherapy.

## Data Availability

The data that support the findings of this study are available from the corresponding author upon reasonable request.

## Author Contributors

All authors provided substantial contributions to the conception (AEO, GB, DM, WDB), design (AEO, DM, ERS), acquisition (AA, MK, MLA, ER), and interpretation (GK, AEO, WB, DM, GB, WDB) of study data and approved the final version of the paper. AEO took the lead in planning the study with support from co-authors. GK carried out the data analysis. AEO is the guarantor.

## Twitter

AEO, GK, AA & IW are in part supported by the National Institute for Health and Care Research (NIHR) Applied Research Collaboration (ARC) Northwest London. The views expressed are those of the authors and not necessarily those of the NHS or the NIHR or the Department of Health and Social Care.

